# Impact of a Population-based Systems Approach on Evidence-based Care for Medicaid-insured Pregnant and Postpartum Women: A Quasi-Experimental Study

**DOI:** 10.1101/2021.03.23.21253829

**Authors:** Lee Anne Roman, Jennifer E. Raffo, Kelly L. Strutz, Zhehui Luo, Melinda Johnson, Peggy VanderMeulen, Susan Henning, Dianna Baker, Claire Titcombe, Cristian I. Meghea

## Abstract

**Introduction:** Evidence-based enhanced prenatal/postnatal care (EPC) programs for Medicaid-insured women have significant positive effects on care and health outcomes. However, EPC enrollment rates are typically low, enrolling less than 30% of eligible women. This study investigated the effects of a population-based systems approach on timely EPC participation and other health care utilization.

**Methods:** This quasi-experimental, population-based, difference-in-difference study used linked birth certificates, Medicaid claims, and EPC data from complete statewide Medicaid birth cohorts retrieved from 2009-2017 and analyzed in 2019-20. System strategies included cross-agency leadership, clinical-community linked practices, Community Health Worker care, mental health coordination, and patient empowerment. Outcomes included EPC participation and early enrollment, prenatal care adequacy, emergency department contact, and postpartum care.

**Results:** Prenatal EPC (7.4 absolute percentage points; 95% CI: 6.3—8.5) and first trimester EPC, (12.4; 95% CI: 10.2—14.5) increased among women served by practices that co-located EPC resources, relative to the comparator group. First trimester EPC improved in the county (17.9; 95% CI: 15.7—20.0); ED decreased in the practices (−11.1; 95% CI: -12.3— -9.9) and postpartum care improved (7.1; 95% CI: 6— 8.2) in the county. EPC participation for Black women served by the practices improved (4.4; 95% CI: 2.2—6.6), as well as early EPC (12.3; 95% CI: 9.0—15.6) and postpartum visits (10.4; 95% CI: 8.3—12.4).

**Conclusions:** A population systems approach improved EPC participation and service utilization for Medicaid-insured women in a county population, for those in practices that co-located EPC resources, and for Black women.

## Introduction

For Medicaid-insured pregnant and postpartum women, select maternal and infant health outcomes have worsened, including maternal morbidity and mortality and preterm birth; with a disproportionate burden for Black women and infants.^1-8^ Moreover, a growing number of beneficiaries enter care with pre-existing or chronic conditions,^9-11^ high levels of stress,^12^ social determinant risk factors^1^, barriers to care^13^, and, for women of color, exposure to racism that influences their health and care;^14-16^ leading to calls for perinatal care improvement.^17-19^

The Centers for Medicare and Medicaid (CMS) Expert Panel on Improving Maternal and Infant Outcomes synthesized best available evidence to improve beneficiary care.^20^ The panel reiterated support for Enhanced Prenatal Care (EPC) programs, available in at least 60% of states, that provide care coordination, social support, linkages to resources, and address social determinants of health.^21^ EPC provides preventive health education and interventions and care coordination, delivered through home visiting, case management, maternity home, doula, navigator or other program models, most with service delivery for women and infants through the postpartum year.^21-24^ Panel recommendations also included developing population-based systems of care to reach and maintain participation of high-risk women, those with complex medical and social problems, in integrated systems of clinical and community-based care.

Rigorous evaluations of state-sponsored Medicaid EPC programs have demonstrated significant risk reduction for adverse birth outcomes and health care improvements.^25-28^ The Michigan Maternal Infant Health Program (MIHP), a federally-designated evidence-based EPC home visiting program, has shown significant reduction of risk for adverse birth outcomes, especially for Black women; improved maternal and infant service use; and reduction of risk for infant mortality.^29-31^ However, statewide EPC programs typically engage less than 30% of eligible women; most women with clinical risk factors do not participate;^32^ and little is known how systems-based recommendations can be accomplished at the population and practice levels to improve timely participation. The objective of this study was to evaluate whether population-based, perinatal system efforts could improve early EPC enrollment and service utilization for Medicaid beneficiaries in a county population in Michigan; for women in high-volume practices that integrate EPC resources; and for Black women, at greater risk for adverse outcomes.

## Methods

### Study Sample

The demonstration county is mixed urban/rural and includes the second-largest city in Michigan, Grand Rapids, with 4,594 births (2009) and about 42% Medicaid births.33 Medicaid eligibility and claims, vital records, and EPC program data were retrieved for complete statewide Medicaid birth cohorts (2009-2017) from the MDHHS Health Services Data Warehouse and were analyzed in 2019-20. Infants’ and mothers’ data, of singleton births, were linked based on unique MDHHS encrypted identifiers, with a linking rate of >95%. The mother-newborn observations were further linked to EPC program data, to Medicaid claims, to supplementary Vital Records data, and to publicly available U.S. Census data. The first baseline year with complete data for all EPC participants was the 2009 birth cohort. Usable baseline data for the practices was not available until 2010. The Michigan State University IRB determined the study did not involve human subjects.

### Population Improvement Activities

All Michigan Medicaid-insured pregnant women are eligible for EPC through Maternal Infant Health Program (MIHP), primarily delivered in the home by nurses and social workers. EPC includes: 1) comprehensive risk screening, 2) standardized interventions based on risks, and 3) care coordination that optimizes resources to address social determinants.^34^ The county has one of several Federal Healthy Start (HS) sites in Michigan where Community Health Workers (CHW), similar in race/ethnicity to their clients, team with EPC providers (EPC+CHW model) to deliver culturally-adapted care and intensive relationship-based care to support client empowerment.^35^ The HS program, Strong Beginnings, engages five health agencies with MIHP programs to deliver team services to Black and Hispanic women, a program with promising results for the reduction of risk for preterm birth.^36^

From 2009-2012, the community engaged in process mapping to discover barriers and gaps in care and underutilization of EPC;^37^ conducted a survey and focus group to identify physician’s knowledge of EPC and referrals for care;^38^ and several agencies initiated agency-specific EPC care improvements. An Agency for Healthcare Research and Quality (AHRQ) award afforded a community planning year (2013), a community infrastructure with a cross-agency physician-led administrative leadership group, multi-agency learning and process improvement efforts, focus groups to elicit patient perspectives,^39^ and a leveraging of administrative and county-level data to guide community improvement efforts that were fully implemented in 2014-15. In addition to community stakeholders, the Michigan Department of Health and Human Services (MDHHS), overseeing the EPC program, helped shape strategies, support data access, and inform policy implications.

A system of care was defined as a spectrum of services that aligns clinical and community resources, builds connections, enhances services that reflect cultural, racial/ethnic and linguistic preferences, and maximizes care coordination for women with complex needs.^40^ The Expanded Chronic Care model, addressing population health, delivery system design, and informed activated patients, was used as an organizing framework.^41^ A key driver diagram, depicts system strategies developed during the AHRQ phase of the demonstration and that were implemented for the next two years. Priority strategies included clinical-community linkages with EPC programs, with on-site resources, in two high volume prenatal practices that serve low-income women. Also, there was community consensus to expand EPC+CHW team care for Black and Hispanic Medicaid-insured women through the local Federal Healthy Start program, prioritizing referrals for women of color, development of CHW patient activation tools and mental health coordination. Detailed information about the community process and improvement strategies are included in an online Perinatal System of Care Toolkit.^42^

### Measures

EPC participation was measured as evidence of any EPC Medicaid claim or program record during pregnancy; early EPC participation was evidence of EPC claims or program record within the first pregnancy trimester. Service utilization indicators from birth certificates and claims included: 1) Kotelchuck adequacy of prenatal care (adequate or adequate plus vs intermediate or inadequate); 2) any prenatal ED use (any ED Medicaid claims during pregnancy vs none); and 3) completion of a postpartum visit in the first 60 days after birth (any postpartum care Medicaid claims vs none in the first 60 days after birth). ACOG recommends a comprehensive postpartum visit no later than 12 weeks after birth.^43^ Postpartum visit measurements were consistent with an Illinois state-wide postpartum visit analyses of Medicaid-insured women which found that a third of women complete a visit less than 21 days,^44^ and to account for the fact that women enrolled in Medicaid on the basis of being pregnant may lose coverage at 60 days postpartum.

Covariates for propensity score estimation included age, education, marital status, an indicator for whether the father was identified on the birth record, maternal and paternal race/ethnicity, maternal alcohol use, tobacco use, prior preterm birth, a previous birth within 18 months of conception, and WIC participation, all from birth records. Medicaid eligibility and claims were used to create an indicator for having Medicaid coverage three months prior to pregnancy and three binary indicators were developed for maternal chronic conditions: asthma (ICD9; 491-493), diabetes (ICD9, 250), and hypertension (ICD9; 401-405). Multiple census variables, both at the block group and census tract level, were used to adjust for poverty and family household characteristics, as well as well-established indexes for community material and social deprivation including Townsend,^45^Jarman,^46^ and Messer^47^ indexes.

### Statistical Analysis

A quasi-experimental pre-post design with a comparison group was used to identify impact of the demonstration. Medicaid-insured women in the county were included in the demonstration and those in the rest of the state formed the pool from which the comparison group was drawn. Analyses were limited to those with singleton births of valid weight and gestational age (≥500 grams and ≥20 weeks) on the birth record. Data were analyzed using difference-in-difference (DID) methodology with period-specific propensity score kernel weighting that accounted for missing covariate data.^48-50^ The DID method ameliorates potential selection biases by subtracting the difference in outcomes between demonstration and comparison groups at the baseline period from the difference in outcomes between demonstration and comparison groups after implementation. The method relies on the assumption that the groups do not systematically change over time, i.e. they would experience the same trend over time had there been no demonstration. However, this assumption would be violated if the women served in the intervention group changed composition, which was likely because the county and intervention sites increased identification of high-risk women. To minimize bias, we used a propensity score-weighted DID method that balances the intervention group at baseline and at follow-up so that the comparison group had similar demographic, geographic, and medical background as the intervention group before and after implementation of strategies.

Outcomes are reported for all women and separately for Black women. Outcomes for women of other races are available in Supplemental materials. The propensity score in the demonstration group was estimated at pre-and post-periods separately, balancing for individual and geographic-level variables. Propensity score kernel weighting was used to estimate the DID effects using linear probability models for each health service outcome. Sensitivity analyses were conducted, and results were qualitatively and quantitatively similar to the main results with all but one estimate having a different direction of the effect (Black women ED use in the practice settings). Stata v.15 (StataCorp, College Station, TX) was used for all analyses.

## Results

Table 1 summarizes characteristics of all Medicaid beneficiaries in the county, the practices, and the state; Table 2 reports characteristics by women who are Black (characteristics of women of other races are in supplemental materials; Appendix 1). Women served by the practices at baseline (n=826) were more likely to be Black (33.9%) or Hispanic (30.0%) compared to the county (n=4,594; 20.5% and 22.7%) and the state of Michigan (n=65,566; 26.9% and 8.3%). They were more likely to have not completed high school (36.7% vs. 29.5% vs 23%), to be enrolled in WIC (74.1% vs. 65.3% vs 69.9%), to have had a prior preterm birth (6.7% vs 2.3% vs. 1.7%), and less likely to be married (21.8% vs. 39.9% vs. 38.2%). Black women, overall, were more likely to have full Medicaid before conception, have chronic health conditions, prior preterm birth and less likely to be married.

**Table 1:** Characteristics of Medicaid-insured Women in Clinical-Community Integrated Practices, County, and Michigan, 2009 and 2015.

|  | Clinical-Community Integrated Practices<sup>a</sup> |  | Demonstration County |  | Michigan Population of Medicaid Births for Comparison Group |  |
| --- | --- | --- | --- | --- | --- | --- |
| Characteristics | 2010<br>N=826 | 2015<br>N=881 | 2009<br>N=4,594 | 2015<br>N=3,937 | 2009<br>N=65,566 | 2015<br>N=58,457 |
| **All women (individual level variable)** | **Mean±SD** | **Mean±SD** | **Mean±SD** | **Mean±SD** | **Mean±SD** | **Mean±SD** |
| Mother's age at birth, years | 24.4±5.6 | 25.7±6.0 | 25.1±5.4 | 26.2±5.6 | 25.1±5.2 | 26.1±5.2 |
| Pre-pregnancy Body Mass In | 27.1±7.2 | 28.5±7.7 | 27.0±6.9 | 28.0±7.2 | 27.1±6.5 | 27.8±7.0 |
|  | **No. (%)** | **No. (%)** | **No. (%)** | **No. (%)** | **No. (%)** | **No. (%)** |
| Black | 280 (33.9) | 304 (34.5) | 940 (20.5) | 952 (24.2) | 15,797 (26.9) | 15,122 (28.4) |
| Hispanic/Latina | 248 (30.0) | 287 (32.6) | 1,042 (22.7) | 829 (21.1) | 4,862 (8.3) | 4,373 (8.2) |
| < High school diploma | 302 (36.7) | 299 (34.0) | 1,355 (29.5) | 899 (22.9) | 13,390 (23.0) | 9,781 (18.5) |
| Married at birth or conception | 180 (21.8) | 226 (25.7) | 1,832 (39.9) | 1,393 (35.4) | 22,455 (38.2) | 18,355 (34.4) |
| Father is Black | 146 (26.8) | 195 (30.2) | 628 (17.5) | 696 (22.2) | 8,948 (20.3) | 9,553 (23.2) |
| Father with < high school diploma | 195 (36.4) | 214 (33.4) | 920 (25.9) | 630 (20.2) | 8,529 (19.5) | 6,713 (16.5) |
| Enrolled in WIC | 612 (74.1) | 633 (72.3) | 2,996 (65.3) | 2,551 (65.6) | 39,001 (69.9) | 36,325 (69.1) |
| Has full Medicaid before conception | 391 (47.3) | 522 (59.3) | 1,400 (30.5) | 1,929 (49.0) | 18,454 (31.4) | 28,588 (53.6) |
| Smoking | 177 (21.4) | 200 (22.7) | 905 (19.7) | 773 (19.6) | 1,7267 (29.6) | 15,391 (29.0) |
| Diabetes prior to/during pregnancy | 45 (5.4) | 50 (5.7) | 216 (4.7) | 210 (5.3) | 2,533 (4.3) | 2,478 (4.6) |
| Hypertension prior to/during pregnancy | 79 (9.6) | 76 (8.6) | 254 (5.5) | 301 (7.6) | 4,704 (8.0) | 4,916 (9.2) |
| Asthma prior to/during pregnancy | 31 (3.8) | 31 (3.5) | 133 (2.9) | 89 (2.3) | 1,809 (3.1) | 1,640 (3.1) |
| Previous preterm birth | 55 (6.7) | 56 (6.4) | 105 (2.3) | 183 (4.7) | 1,010 (1.7) | 1,710 (3.2) |
| Short interpregnancy interval<sup>b</sup> | 191 (23.1) | 225 (25.5) | 1,137 (24.7) | 969 (24.6) | 13,845 (23.6) | 13,440 (25.2) |
| **All women (Census tract level variable)** | **Mean±SD** | **Mean±SD** | **Mean±SD** | **Mean±SD** | **Mean±SD** | **Mean±SD** |
| % Families < FPL | 21.1±12.9 | 22.8±14.9 | 15.4±11.2 | 18.2±13.8 | 16.0±13.4 | 20.0±15.3 |
| % Female head of household | 29.4±14.2 | 32.1±14.3 | 24.5±12.2 | 28.2±13.7 | 25.6±17.5 | 28.2±17.6 |
| % Families with some college, associate degree | 32.7±7.4 | 33.4±10.2 | 32.4±8.1 | 34.5±9.2 | 34.1±8.7 | 36.9±8.8 |
| Townsend index | 1.8±2.9 | 2.4±2.7 | 0.8±2.7 | 1.4±2.9 | 1.0±2.8 | 1.2±2.9 |
| Jarman index | 4.3±5.3 | 4.7±5.3 | 2.9±5.0 | 3.3±5.1 | 1.7±4.0 | 1.9±4.1 |

Abbreviations: SD, standard deviation; BMI, body mass index; WIC, Special Supplemental Nutrition Program for Women, Infants, and Children; FPL, federal poverty level
<sup>a</sup> 2010-2015 due to incomplete data in 2009
<sup>b</sup> Less than 18 months from previous birth to conception

**Table 2:** Characteristics of Black Medicaid-insured Women in Clinical-Community Integrated Practices, County, and Michigan, 2009 and 2015.

| Characteristics | Clinical-community integrated Practices<sup>a</sup> |  | Demonstration County |  | Michigan population of Medicaid births for comparison group |  |
| --- | --- | --- | --- | --- | --- | --- |
|  | 2010<br>N=280 | 2015<br>N=304 | 2009<br>N=940 | 2015<br>N=952 | 2009<br>N=15,797 | 2015<br>N=15,122 |
| **All women (individual level variable)** | **Mean±SD** | **Mean±SD** | **Mean±SD** | **Mean±SD** | **Mean±SD** | **Mean±SD** |
| Mother's age at birth, years | 23.8±5.3 | 24.9±5.7 | 24.1±5.6 | 25.5±5.5 | 24.3±5.7 | 25.4±5.4 |
| Pre-pregnancy body mass index (BMI) | 27.9±7.7 | 29.2±8.1 | 28.3±7.7 | 29.4±8.1 | 27.9±7.2 | 28.8±7.8 |
|  | **No. (%)** | **No. (%)** | **No. (%)** | **No. (%)** | **No. (%)** | **No. (%)** |
| Hispanic/Latina | 10 (3.6) | 12 (3.9) | 24 (2.6) | 28 (2.9) | 230 (1.5) | 246 (1.6) |
| < High school diploma | 100 (35.8) | 81 (26.7) | 290 (30.9) | 232 (24.4) | 4,367 (28.1) | 3,043 (20.4) |
| Married at birth or conception | 34 (12.1) | 36 (11.8) | 163 (17.3) | 151 (15.9) | 2,248 (14.2) | 2,035 (13.5) |
| Father is Black | 110 (85.9) | 155 (87.1) | 447 (88.5) | 493 (85.6) | 7,226 (94.3) | 7,748 (92.8) |
| Father with < high school diploma | 31 (24.4) | 34 (19.2) | 111 (22.6) | 90 (15.7) | 1,384 (18.5) | 1,265 (15.5) |
| Enrolled in WIC | 206 (73.6) | 232 (76.6) | 703 (74.9) | 713 (75.4) | 10,791 (75.3) | 11,044 (74.3) |
| Has full Medicaid before conception | 187 (66.8) | 207 (68.1) | 511 (54.4) | 625 (65.7) | 7,377 (46.7) | 10,054 (66.5) |
| Smoking | 57 (20.4) | 64 (21.1) | 171 (18.2) | 164 (17.2) | 2,912 (18.6) | 2,942 (19.6) |
| Diabetes prior to/during pregnancy | 14 (5.0) | 13 (4.3) | 54 (5.7) | 47 (4.9) | 684 (4.3) | 762 (5.0) |
| Hypertension prior to/during pregnancy | 32 (11.4) | 45 (14.8) | 89 (9.5) | 114 (12.0) | 1,586 (10.0) | 1,779 (11.8) |
| Asthma prior to/during pregnancy | 12 (4.3) | 12 (3.9) | 43 (4.6) | 30 (3.2) | 651 (4.1) | 647 (4.3) |
| Previous preterm birth | 25 (8.9) | 35 (11.5) | 38 (4.0) | 77 (8.1) | 255 (1.7) | 546 (3.6) |
| Short interpregnancy interval<sup>b</sup> | 74 (26.4) | 85 (28.0) | 243 (25.9) | 247 (25.9) | 3,688 (23.3) | 4,023 (26.6) |
| **All women (Census tract level variable)** | **Mean±SD** | **Mean±SD** | **Mean±SD** | **Mean±SD** | **Mean±SD** | **Mean±SD** |
| % Families < FPL | 23.9±11.9 | 24.4±14.9 | 20.3±10.5 | 22.3±14.2 | 26.0±15.5 | 30.8±16.1 |
| % Female head of household | 35.9±14.1 | 36.7±14.6 | 32.6±12.2 | 34.7±14.1 | 44.0±18.1 | 45.9±16.9 |
| % Families with some college, associate degree | 32.7±7.3 | 34.5±10.3 | 32.1±7.8 | 34.9±9.8 | 35.1±9.6 | 38.0±9.5 |
| Townsend index | 2.7±2.7 | 2.8±2.4 | 2.1±2.4 | 2.5±2.5 | 3.1±2.6 | 3.3±2.5 |
| Jarman index | 4.9±4.9 | 4.8±4.7 | 4.5±4.5 | 4.7±4.6 | 3.8±3.7 | 3.9±3.5 |

Abbreviations: SD, standard deviation; BMI, body mass index; WIC, Special Supplemental Nutrition Program for Women, Infants, and Children; FPL, federal poverty level
<sup>a</sup> 2010-2015 due to incomplete data in 2009
<sup>b</sup> Less than 18 months from previous birth to conception

### County Results

In the unadjusted pre-post demonstration comparison (Table 3), there were larger improvements in first trimester EPC enrollment in the county (17.5 APP) vs. statewide (3.4 APP). Black women had larger pre-post demonstration improvements in first-trimester EPC enrollment in the county (18 APP) vs. statewide (4.2 APP). However, the state-wide comparator group had modest increases in EPC participation statewide vs. the county (−1.6 APP). Propensity score-weighted DID findings (Table 3, adjusted DID) showed large increases in first trimester EPC enrollment among all women served in the county (17.9 APP; 95% CI: 15.7—20.0) and among Black women in the county (12.3 APP; 95% CI: 9.0—15.6).

**Table 3:** Propensity Score Weighted Difference-in-Different Estimates on EPC and Health Service Outcomes among Medicaid-Insured Women.

|  | Clinical-Community Integrated Practices<sup>a</sup> |  | Demonstration County |  | Michigan population of Medicaid births for comparison group |  | Clinical-Community Integrated Practices vs. State Comparators | Demonstration County vs. State Comparators |
| --- | --- | --- | --- | --- | --- | --- | --- | --- |
|  | 2010 | 2015 | 2009 | 2015 | 2009 | 2015 | DID | DID |
|  | Unadjusted %<br>N=826 | Unadjusted %<br>N=881 | Unadjusted %<br>N=4,594 | Unadjusted %<br>N=3,937 | Unadjusted %<br>N=65,566 | Unadjusted %<br>N=58,457 | Adjusted APP | Adjusted APP |
| **All women** |  |  |  |  |  |  |  |  |
| EPC participation | 58.0 | 66.3 | 32.1 | 34.2 | 23.9 | 27.5 | 7.4**<br>(6.3 – 8.5) | -1.6**<br>(-2.6 – -0.6) |
| First trimester EPC<sup>b</sup> | 54.5 | 67.8 | 35.1 | 52.6 | 33.9 | 37.3 | 12.4**<br>(10.2 – 14.5) | 17.9**<br>(15.7 – 20.0) |
| Adequate Prenatal Care | 67.4 | 77.1 | 74.0 | 77.4 | 75.0 | 76.2 | 3.2**<br>(2.1 – 4.3) | 2.2**<br>(1.2 – 3.2) |
| Any ED use during pregnancy | 56.1 | 54.6 | 29.7 | 41.2 | 37.9 | 49.3 | -11.1**<br>(-12.3 – -9.9) | -0.6<br>(-1.7 – 0.5) |
| Postpartum care within 60 days | 82.2 | 81.6 | 59.1 | 68.9 | 54.2 | 58.7 | 0.4<br>(-0.6 – 1.4) | 7.1**<br>(6.0 – 8.2) |
| **Black women** | **N=280** | **N=304** | **N=940** | **N=952** | **N=15,797** | **N=15,122** |  |  |
| EPC participation | 63.9 | 68.4 | 44.0 | 48.5 | 32.4 | 43.1 | 4.4**<br>(2.2 – 6.6) | -0.1<br>(-2.2 – 2.0) |
| First trimester EPC | 48.0 | 56.3 | 30.9 | 48.9 | 25.2 | 29.4 | 3.3<br>(-0.1 – 6.7) | 12.3**<br>(9.0 – 15.6) |
| Adequate prenatal care | 67.6 | 71.1 | 74.7 | 76.0 | 74.8 | 76.3 | -10.0**<br>(-12.1 – -7.9) | -0.7<br>(-2.6 – 1.2) |
| Any ED use during pregnancy | 67.9 | 66.8 | 49.4 | 60.9 | 53.8 | 68.3 | -6.5**<br>(-8.6 – -4.3) | 2.5*<br>(0.3 – 4.6) |
| Postpartum care within 60 days | 77.5 | 81.9 | 64.0 | 79.5 | 52.0 | 58.8 | 1.8<br>(-0.2 – 3.8) | 10.4**<br>(8.3 – 12.4) |

Abbreviations: DID, difference-in-difference; APP, Absolute Percentage Points; EPC, enhanced prenatal care; ED, emergency department.
<sup>a</sup> 2010 data used as pre-demonstration period due to incomplete data in 2009, <sup>b</sup> Among EPC participants, \* p-value < 0.05, \*\* p-value < 0.01

Table 3 reports larger unadjusted pre-post improvements in the share of pregnancies with adequate prenatal care among all women in the county (3.4 APP) vs. the statewide comparison group (1.2 APP). There were pre-post increases in the county in the share of women with postpartum care within 60 days after birth (9.8 APP) vs. statewide (4.5 APP). Black women also experienced larger pre-post improvements in the likelihood of receiving postpartum care in the first 60 days after birth in the county (15.5 APP) vs. statewide (6.8 APP).

There were modest propensity score-weighted DID effects (Table 3, adjusted DID) of the demonstration increasing the share of women with adequate or better prenatal care for all women in the county (2.2 APP; 95% CI: 1.2—3.2). There was relatively large DID effects of the demonstration in the county increasing the share of women with appropriate postpartum care (7.1 APP; 95% CI: 6.0—8.2) and among Black women (10.4 APP; 95% CI: 8.3—12.4).

### Clinical-Community Integrated Practices Results

In the unadjusted pre-post demonstration comparison (Table 3), there were larger absolute percentage points (APP) improvements between 2009-2015 in EPC participation among all women served in the integrated practices (8.3 APP) vs. the statewide comparison group (3.6 APP), and larger improvements in first trimester EPC enrollment in the integrated practices (13.3 APP) vs. statewide (3.4 APP). Black women had larger pre-post demonstration improvements in first-trimester EPC enrollment when served by the integrated practices (8.3 APP) vs. statewide (4.2 APP).

Propensity score-weighted DID findings (Table 3, adjusted DID) indicate demonstration effects increasing EPC participation among all women served in the integrated practices (7.4 APP; 95% CI: 6.3— 8.5) and for Black women (4.4 APP; 95% CI: 2.2—6.6). There were large increases in first trimester EPC enrollment among all women served by the practices (12.4 APP; 95% CI: 10.2—14.5).

Table 3 also reports larger unadjusted pre-post improvements in the share of pregnancies with adequate prenatal care among all women in the integrated practices (9.7 APP) vs. the statewide comparison group (1.2 APP). There were modest propensity score-weighted DID effects (Table 3, adjusted DID) of the demonstration increasing the share of women with adequate or better prenatal care for all women served by the integrated practices (3.2 APP; 95% CI: 2.1—4.3). There were relatively large DID effects of the demonstration reducing the share of all women with an ED visit during pregnancy when served by the integrated practices (−11.1 APP; 95% CI: -12.3— -9.9), Black women (−6.5 APP; 95% CI: -8.6— -4.3).

## Discussion

In the context of persistent disparities and underutilization of services, implementation of population-based system strategies improved overall EPC enrollment for women served by practices with clinical-community EPC linkages for Medicaid-insured pregnant women. Large impacts were also seen in first trimester EPC screening, both in the county population and among the women served by the integrated practices. Improvements were noted for adequacy of prenatal care, in the county population and in the practice group; ED utilization in the practice group; and postpartum care improvements countywide. Benefits for Black women included greater EPC participation in the practice settings; and first trimester EPC enrollment and postpartum care for the county.

To put improvements in perspective, the increases in first trimester EPC enrollment represent over 50% relative improvement from the baseline level for the county, with early enrollment of more than half of all EPC participants. Early enrollment findings are important for first trimester risk assessment, connections to community resources, and initiation of EPC interventions.^51^ EPC enrollments in the linked practices reached 66% for all and 68% for Black women; however, the lack of improvements in county EPC participation was not entirely unexpected. After an early increase in EPC enrollment (32% to 39%), a loss of revenue for the EPC-CHW model temporarily reduced capacity, with re-building of caseloads during 2014-2015.

For Black women living in the county, the 40% relative improvement in first trimester EPC, resulting in 48.9% of all Black women enrolling early is notable. There was significant improvement for Black women served by the integrated practices for overall EPC enrollment (7%). For postpartum care, the relative county improvements were larger for Black women (16%) than women of other races (11%). While there are several studies focused on system of care approaches, some targeting geographic populations;^51-54^ we are not aware of comparative studies of system of care and EPC, early enrollment, and service utilization using DID propensity score methods over extended periods of time.

Our results are novel as few studies have examined how population approaches can improve the delivery of evidence-based, community-delivered programs and health service utilization. Further, findings inform the calls for systems change to address population health care, the redesign of care linking clinical-community providers, and for reducing socioeconomic and racial/ethnic disparities in Medicaid-insured populations.^55-60^ For clinicians, directed to address social determinants, EPC and CHW providers can be important sources of preventive health education and interventions, health monitoring, connections to resources, and support during pregnancy through the postpartum transition to well woman primary care, especially for those with chronic conditions. The postpartum visit improvements for Black women in the overall county are especially important with current efforts to address maternal morbidity.^61^

However, the demonstration underscored the challenge of increasing community EPC and CHW capacity given current reimbursement mechanisms, with reported uncompensated EPC program costs up to 40%, and few stable funding mechanisms for CHW providers delivering race-concordant care for low-income women of color.^62^ Policymakers, health plans, public health, health systems, and clinicians, with resource constraints, need testing of innovative delivery system redesign and payment models to reduce maternal socioeconomic and racial/ethnic disparities.^63, 64^ For example, MDHHS state policymakers and the local federal HS program are engaged in a five-year demonstration of a Pay-for-Success financial model for the EPC-CHW team intervention targeting reduction of preterm birth and rapid repeat pregnancy.^65^ This effort has allowed for continuous improvement efforts, tracking of implementation and health care indicators reported here, as well as, evaluation of health outcomes. A study of multi-level interventions to address maternal morbidity and mortality in Black women is also being initiated in the demonstration county (Kent County/Grand Rapids) and in Genesee County/Flint to expand EPC telehealth approaches to address patient preferences and increase EPC participation.^66^ These communities are also pilot sites for the national AIM – Community Care Initiative with efforts to develop systemic processes to assist women in connecting to community-based services in outpatient and community settings.^67^

### Limitations

The strength of this study was the sustained engagement of community stakeholders who delivered multiple improvement strategies, with potentially synergistic effects, overtime, and the use of existing administrative data. However, while difference-in-difference methodology can be used to estimate causal effect, future research is needed to isolate the most relevant components of system-of-care interventions on care and health outcomes, including maternal morbidity and birth outcomes. Matching was restricted to observable characteristics and the use of administrative data has limitations with potential inaccuracies for medical risk factors. Last, although systems of care approaches share common features, system strategies, by definition, are tailored to the community.

## Conclusions

Population-based system strategies improved participation in an evidence-based, community-delivered EPC program and other perinatal service utilization relevant for the maternal and child health of Medicaid beneficiaries. Establishing clinical-community EPC linkages in high-volume practices were especially effective in increasing EPC enrollment for all patients and for Black women.

## Data Availability

Data was used through a data use agreement with the Michigan Department of Health and Human Services that does not allow us to share data.

## Acknowledgements

Community stakeholders shared project leadership (Drs. Stephen Rechner and Richard Leach) and action work groups; implemented strategies; and informed interpretation of analyses (Spectrum Health, Cherry Health, Kent County Health Department, Arbor Circle). Maternal Infant Health Program staff and Strong Beginnings equity and education coordinators, Community Health Workers, and program participants informed the development of system of care strategies and/or delivered services. The Michigan Department of Health and Human Services (MDHHS) provided access to the Health Services Data Warehouse; the Director of the Division of Maternal and Infant Health, Dawn Shanafelt, sponsored the project; and staff participated in leadership meetings, works groups, and informed policy. This project was supported by grant number R18HS020208 from the Agency for Healthcare Research and Quality. The content is solely the responsibility of the authors and does not necessarily represent the official views of the Agency for Healthcare Research and Quality.

## Appendix Table

**Table 1:**
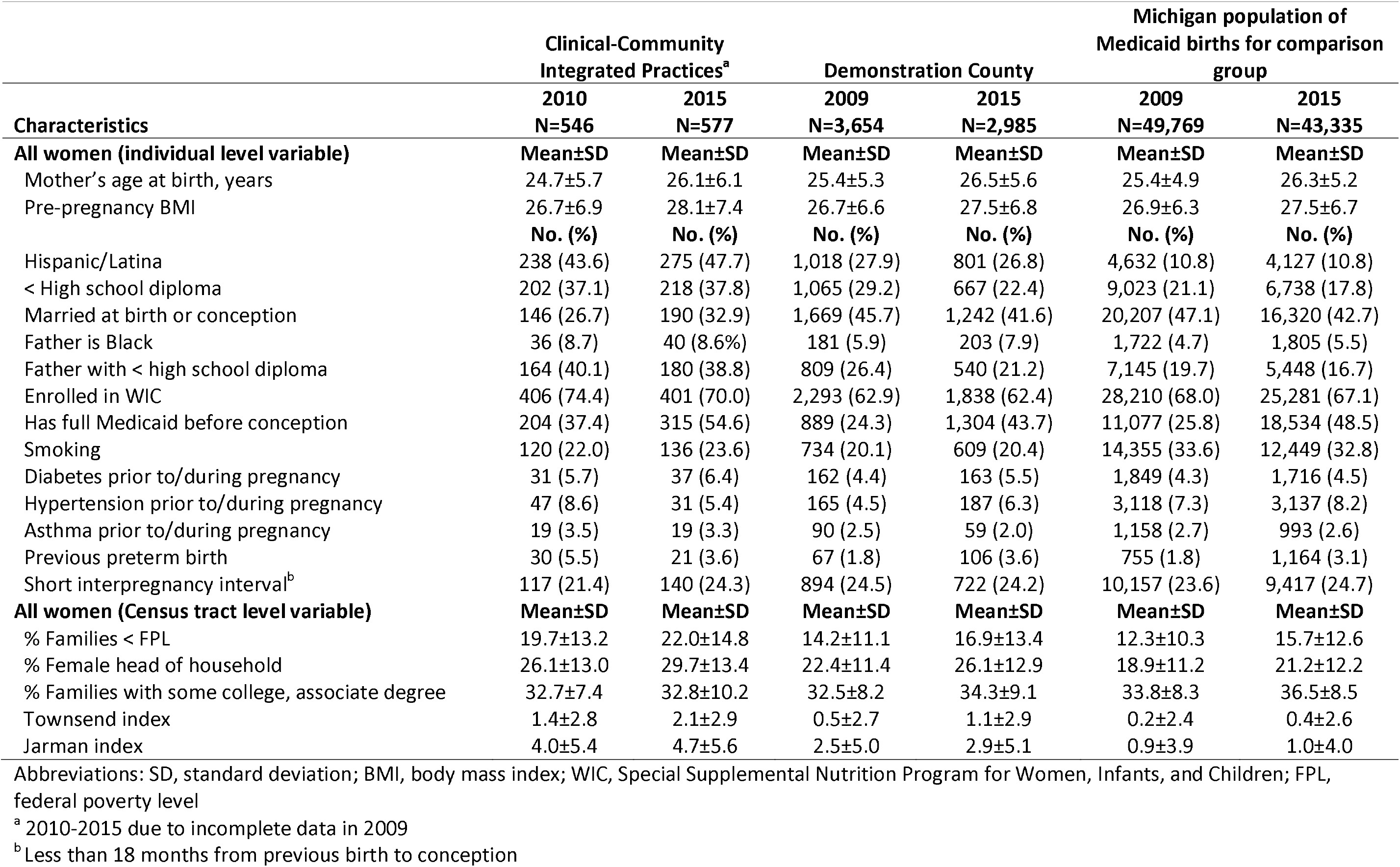
Characteristics of Black Medicaid-insured Women in Clinical-Community Integrated Practices, County, and Michigan, 2009 and 2015.

| Characteristics | Clinical-Community Integrated Practices<sup>a</sup> |  | Demonstration County |  | Michigan population of Medicaid births for comparison group |  |
| --- | --- | --- | --- | --- | --- | --- |
|  | 2010 | 2015 | 2009 | 2015 | 2009 | 2015 |
|  | N=546 | N=577 | N=3,654 | N=2,985 | N=49,769 | N=43,335 |
| **All women (individual level variable)** | **Mean±SD** | **Mean±SD** | **Mean±SD** | **Mean±SD** | **Mean±SD** | **Mean±SD** |
| Mother's age at birth, years | 24.7±5.7 | 26.1±6.1 | 25.4±5.3 | 26.5±5.6 | 25.4±4.9 | 26.3±5.2 |
| Pre-pregnancy BMI | 26.7±6.9 | 28.1±7.4 | 26.7±6.6 | 27.5±6.8 | 26.9±6.3 | 27.5±6.7 |
|  | **No. (%)** | **No. (%)** | **No. (%)** | **No. (%)** | **No. (%)** | **No. (%)** |
| Hispanic/Latina | 238 (43.6) | 275 (47.7) | 1,018 (27.9) | 801 (26.8) | 4,632 (10.8) | 4,127 (10.8) |
| < High school diploma | 202 (37.1) | 218 (37.8) | 1,065 (29.2) | 667 (22.4) | 9,023 (21.1) | 6,738 (17.8) |
| Married at birth or conception | 146 (26.7) | 190 (32.9) | 1,669 (45.7) | 1,242 (41.6) | 20,207 (47.1) | 16,320 (42.7) |
| Father is Black | 36 (8.7) | 40 (8.6%) | 181 (5.9) | 203 (7.9) | 1,722 (4.7) | 1,805 (5.5) |
| Father with < high school diploma | 164 (40.1) | 180 (38.8) | 809 (26.4) | 540 (21.2) | 7,145 (19.7) | 5,448 (16.7) |
| Enrolled in WIC | 406 (74.4) | 401 (70.0) | 2,293 (62.9) | 1,838 (62.4) | 28,210 (68.0) | 25,281 (67.1) |
| Has full Medicaid before conception | 204 (37.4) | 315 (54.6) | 889 (24.3) | 1,304 (43.7) | 11,077 (25.8) | 18,534 (48.5) |
| Smoking | 120 (22.0) | 136 (23.6) | 734 (20.1) | 609 (20.4) | 14,355 (33.6) | 12,449 (32.8) |
| Diabetes prior to/during pregnancy | 31 (5.7) | 37 (6.4) | 162 (4.4) | 163 (5.5) | 1,849 (4.3) | 1,716 (4.5) |
| Hypertension prior to/during pregnancy | 47 (8.6) | 31 (5.4) | 165 (4.5) | 187 (6.3) | 3,118 (7.3) | 3,137 (8.2) |
| Asthma prior to/during pregnancy | 19 (3.5) | 19 (3.3) | 90 (2.5) | 59 (2.0) | 1,158 (2.7) | 993 (2.6) |
| Previous preterm birth | 30 (5.5) | 21 (3.6) | 67 (1.8) | 106 (3.6) | 755 (1.8) | 1,164 (3.1) |
| Short interpregnancy interval<sup>b</sup> | 117 (21.4) | 140 (24.3) | 894 (24.5) | 722 (24.2) | 10,157 (23.6) | 9,417 (24.7) |
| **All women (Census tract level variable)** | **Mean±SD** | **Mean±SD** | **Mean±SD** | **Mean±SD** | **Mean±SD** | **Mean±SD** |
| % Families < FPL | 19.7±13.2 | 22.0±14.8 | 14.2±11.1 | 16.9±13.4 | 12.3±10.3 | 15.7±12.6 |
| % Female head of household | 26.1±13.0 | 29.7±13.4 | 22.4±11.4 | 26.1±12.9 | 18.9±11.2 | 21.2±12.2 |
| % Families with some college, associate degree | 32.7±7.4 | 32.8±10.2 | 32.5±8.2 | 34.3±9.1 | 33.8±8.3 | 36.5±8.5 |
| Townsend index | 1.4±2.8 | 2.1±2.9 | 0.5±2.7 | 1.1±2.9 | 0.2±2.4 | 0.4±2.6 |
| Jarman index | 4.0±5.4 | 4.7±5.6 | 2.5±5.0 | 2.9±5.1 | 0.9±3.9 | 1.0±4.0 |

Abbreviations: SD, standard deviation; BMI, body mass index; WIC, Special Supplemental Nutrition Program for Women, Infants, and Children; FPL, federal poverty level
<sup>a</sup> 2010-2015 due to incomplete data in 2009
<sup>b</sup> Less than 18 months from previous birth to conception

